# Adjunct tirofiban treatment after successful endovascular thrombectomy recanalisation in acute anterior circulation ischemic stroke (ATTRACTION): protocol of a multicenter, prospective, double-blind, randomised trial

**DOI:** 10.64898/2026.02.25.26346143

**Authors:** Xiang Luo, Hao Huang, Shabei Xu, Guo Li, Yi Zhang, Yiming Luo, Qianqian Kong, Yi Xie, Chenchen Liu, Gang Deng, Yihui Wang, Donghui Ao, Lingning Lan, Ying Yu, Zhouping Tang, Wei Wang

**Author notes:** **Correspondence to**, Wei Wang, Address: No.1095, Jiefang Avenue, Qiaokou District, Wuhan, 430030, China., Zhouping Tang, Address: No.1095, Jiefang Avenue, Qiaokou District, Wuhan, 430030, China. Xiang Luo, Address: No.1095, Jiefang Avenue, Qiaokou District, Wuhan, 430030, China. Xiang Luo,Hao Huang and Shabei Xu contributed equally.

## Abstract

**Background:** Successful recanalisation without functional independence is a frequent phenomenon following endovascular thrombectomy for large vessel occlusion stroke.

**Aim:** To demonstrate safety and efficacy of adjunct tirofiban therapy after endovascular thrombectomy in patients with anterior circulation large vessel occlusion stroke achieving successful recanalization defined as modified Thrombolysis In Cerebral Infarction (mTICI) 2b-3.

**Design:** The study of adjunct tirofiban treatment after successful endovascular thrombectomy recanalisation (ATTRACTION) is a multicenter, prospective, double-blind, randomized trial enrolling 1360 patients in China. Eligible patients will be randomised 1:1 to either the tirofiban or placebo group.

**Outcome:** The primary efficacy outcomes is assessed as the proportion of participants with a modified Rankin Scale (mRS) score of 0-2 at 90 days, and the primary safety outcome is symptomatic intracranial haemorrhage within 48 hours from randomisation.

**Conclusion:** This study will provide evidence on the efficacy and safety of sequential tirofiban therapy after successful recanalisation in patients with anterior circulation large vessel occlusion stroke.

**Trial registration number:** NCT06265051

**WHAT IS ALREADY KNOWN ON THIS TOPIC:** Successful recanalization without functional independence is a frequent phenomenon following endovascular thrombectomy and previous small-sample, retrospective studies supported the administration of adjunct tirofiban therapy in patients after endovascular thrombectomy achieving successful recanalization.

**WHAT THIS STUDY ADDS:** The ATTRACTION trial aims to access the efficacy and safety of adjunct tirofiban therapy and the protocol describes the rationale and design of the trial.

**HOW THIS STUDY MIGHT AFFECT RESEARCH, PRACTICE OR POLICY:** ATTRACTION trial will inform whether tirofiban therapy after successful recanalisation by endovascular thrombectomy can improve patient outcomes.

## INTRODUCTION

Endovascular thrombectomy (EVT) is the established standard therapy for acute ischemic stroke due to anterior circulation large vessel occlusion (LVO). Despite achieving successful recanalization in > 70% of patients, only 40%-50% attain functional independence at 90 days.^[1]^ This discrepancy underscores multifactorial reperfusion failure, driven by irreversible infarct evolution at intervention, microcirculatory dysfunction, comorbid vulnerabilities, and post-stroke complications.^[2,3]^

Adjunctive pharmacotherapy to mitigate these mechanisms remains an unmet need. While the CHOICE trial reported improved outcomes with intra-arterial alteplase after EVT-mediated recanalisation, ^[4]^ subsequent trials (POST-UK, ^[5]^ POST-TNK, ^[6]^ ANGEL-TNK, ^[7]^ ATTENTION IA ^[8]^) yielded conflicting results. Although a recent meta-analysis demonstrated that adjunct intra-arterial thrombolysis significantly increased the rate of favorable functional outcome, it also highlighted substantial heterogeneity among the included studies, underscoring the need for further validation in larger, well-designed trials ^[9]^ These inconsistencies, compounded by thrombolytic contraindications and bleeding risks, limit broad clinical applicability.

Tirofiban, a small-molecule non-peptide tyrosine derivative, acts as a selective, competitive, dose-dependent, and reversible inhibitor of the glycoprotein (GP) IIb/IIIa receptor. It blocks fibrinogen binding to platelets and subsequent process of platelet aggregation. ^[10-13]^ It was originally applied for ischemic heart disease, demonstrating both efficacy and acceptable tolerability.^[12]^ Building on the theoretical foundation of tirofiban’s use in ischemic heart disease, neurologists have investigated its applicability in acute ischemic stroke. Recent studies have validated the safety of tirofiban in stroke patients who underwent EVT.^[15]^ Distinct from fibrinolytic agents, GP IIb/IIIa inhibitors possess no thrombolytic properties. Instead, tirofiban works by blocking the final common pathway of activated platelet aggregation and subsequent thrombus formation, thereby aiming to inhibit thrombosis secondary to endothelial injury and prevent re-occlusion. ^[13]^ Although emerging evidence supports its therapeutic role following EVT for large vessel occlusion stroke,^[14,15]^ robust evidence from randomized controlled trials evaluating adjunct tirofiban after successful recanalization in acute ischemic stroke remains scarce.

To address this gap, we initiated the Adjunct Tirofiban Treatment after Successful Endovascular Thrombectomy Recanalisation in Acute Anterior Circulation Ischemic Stroke (ATTRACTION) trial aiming to determine whether adjunct tirofiban improves outcomes in Chinese stroke patients achieving successful recanalization after EVT.

## METHODS

### Design

The ATTRACTION trial is an investigator-initiated, multicenter, prospective, randomized, double-blind trial in China, and its primary objective is to assess whether tirofiban can improve neurological outcomes in patients with acute anterior circulation LVO stroke who reach successful recanalization with EVT within 24 hours of symptom onset. The ATTRACTION trial is registered on www.clinicaltrials.gov (NCT06265051) on February 2024. The study design is shown in figure 1. A detailed schedule is depicted in Supplementary Figure 1.

**Figure 1.**
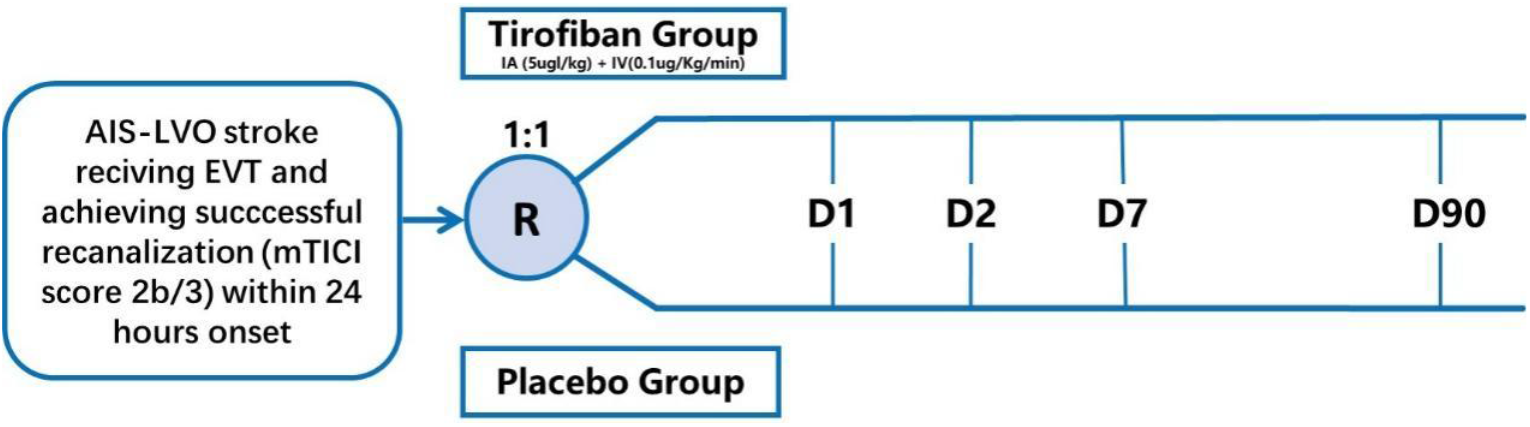
Study design. AIS, acute ischemic stroke; EVT, endovascular thrombectomy, LVO, large-vessel occlusion;

### Study population

Patients are enrolled from about 80 clinical centers across China (Supplementary Figure 2 for geographic distribution) who recruit adults with acute LVO stroke who have achieved successful recanalisation after EVT, starting in April 2024. Eligible patients will be systematically enrolled by investigators following standardized screening procedures. The inclusion and exclusion criteria are summarized in figure 2. Notably, enrollment included patients with tandem occlusions. Furthermore, patients based on prior administration of standard intravenous thrombolysis before EVT were also eligible.

**Figure 2.**
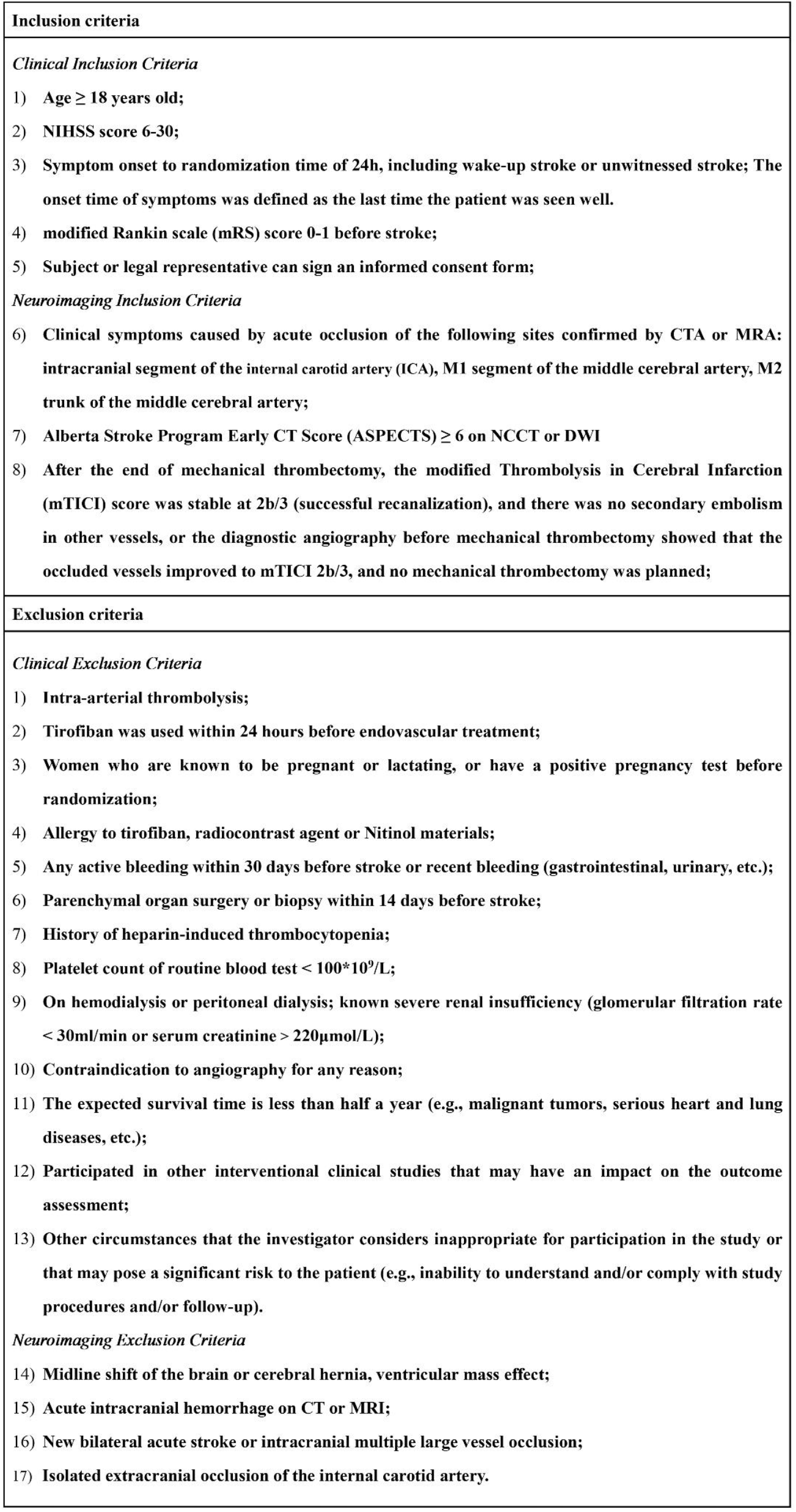
Inclusion and exclusion criteria.

### Randomisation

The eligible patients will be randomised to treatment with either tirofiban or placebo with a ratio of 1:1 via a 24/7 real-time central network system according to the block randomization method with block size of 4. Randomisation is stratified according to clinical centers. The study is double-blind, and neither the investigators nor the experimentalists are aware of the treatment assignments. Unblinding is necessary when needed.

### Intervention

Both treatment group will undergo endovascular treatment. The EVT procedures will be performed according to the routine of each center with devices approved by the National Medical Products Administration. Endovascular treatment consisted of thrombus aspiration, mechanical thrombectomy, or various combinations of these approaches. Patients undergoing remedial treatments such as balloon angioplasty or stent placement are still included in the study but not those receiving intra-arterial thrombolysis (including intra-arterial tirofiban). After the endovascular treatment, cases are evaluated based on the final reperfusion mTICI score for inclusion. Participants achieving successful recanalization (mTICI 2b-3) are randomised to receive either tirofiban or matched placebo through double-blinded allocation, with blinded study drug infusion initiated immediately following randomisation. The tirofiban treatment is intra-arterial tirofiban (Xinweining: 100ml/5mg tirofiban) administration at 5ug/kg(0.1ml/kg) bolus (not exceeding 0.5 mg) at a rate of 1 ml/min, subsequently continued as intravenous infusion at 0.1ug/kg/min (0.12ml/kg/h) for up to 24 hours. ^[11,15]^ The placebo consists of normal saline, administered using the same infusion volumes and protocol as the tirofiban solution. The tirofiban and placebo have the same appearance and packaging to ensure our double-blindness.

### Efficacy outcomes

The primary efficacy outcome is assessed as the proportion of patients with a modified Rankin Scale (mRS) score of 0-2 at 90 (±7) days. Secondary efficacy outcomes comprise the proportion with mRS score 0-1 at 90 (±7) days, the proportion with mRS score 0-3 at 90 (±7) days, the distribution of mRS score (shift analysis) at 90 (±7) days, the proportion with NIHSS score 0-1 or ≥ 4-point reduction from baseline at 36 (±12) hours, and the EQ-5D-5L score at 90 (±7) days. Outcome assessment was centrally adjudicated based on audiovisual records by certified blinded raters. In the absence of such recordings, score assessment was determined by locally certified investigators blinded to the treatment assignment. All mRS assessors underwent specific training.

### Safety outcomes

The primary safety outcome is defined as the proportion of symptomatic intracranial haemorrhage (sICH) as per Heidelberg Bleeding Classification within 48 hours after randomisation.^[16]^ Furthermore, the secondary safety outcomes include all-cause mortality at 90 (±7) days and the proportion of any intracranial haemorrhage (ICH) within 48 hours after randomisation.

### Sample size

This study is a randomised controlled trial design, whose primary efficacy outcome measure is 90 (±7) days mRS score 0-2 proportion. According to the subgroup analysis of the RESCUE BT study investigating the effect of adjunct intravenous tirofiban therapy following successful reperfusion, the placebo group exhibited a 44.0% rate of mRS 0-2, compared to 54.3% in the tirofiban group.^[9]^ In this trial, the assumed mRS 0-2 rate was set at 44.0% for the placebo group with an 8% improvement (a more conservative between-group difference assumption) in the tirofiban group. The sample size ratio between the treatment and control arm is 1:1. In order to demonstrate the expected treatment effect with a 5% two-sided type I error, 80% statistical power and 10% loss of follow-up, the required sample size is 1360 with 680 participants in each group in this trial.

### Data management and quality control

This study employed an Electronic Data Capture (EDC) system for data collection and entry. We ensured data quality by using methods such as double entry and range checks. The Data and Safety Monitoring Board, which consists of a neurologist, a neurointerventionalist and a statistician independent from the study, oversees the conduct of the trial and monitors patient safety by reviewing all serious adverse events. Adverse events were evaluated using the 5th edition of the CTCAE (Common Terminology Criteria for Adverse Events) of the United States.^[17]^ Based on identified safety concerns, the DSMB holds the authority to recommend the continuation, modification, or termination of the trial to the Steering Committee.

### Statistical analysis

All analyses will be conducted using R (version 4.3.0; R Foundation for Statistical Computing, Vienna, Austria) and SAS (version 9.4; SAS Institute Inc., Cary, NC) following the pre-specified statistical analysis plan whose completion is scheduled to be finalized before the locking of the database and the unblinding of the study. The Full Analysis Set (FAS) will comprise all randomised participants who received either medical therapy or endovascular therapy (EVT), adhering to the intention-to-treat (ITT) principle. A Per-Protocol Set (PPS) will be defined as participants completing protocol-specified treatments without major deviations. For baseline characteristics, continuous variables will be analyzed using Wilcoxon rank-sum test or Student’s t-test, while categorical variables will be assessed using Fisher’s exact test, χ^2^ test, or Wilcoxon rank-sum test as appropriate. Primary efficacy outcomes will be assessed in the FAS and PPS using modified Poisson regression model to estimate risk ratios (RR) with 95% confidence intervals (CI). Secondary outcomes analysis methods include: modified Poisson regression model for the other binary data and Win Ratio method for the EQ-5D-5L. For the ordinal mRS, ordinal logistic regression model will be used to test for a difference between treatments, with common odds ratio and corresponded 95%CI being reported. Brant test will be used to assess the proportional odds assumption. If the proportional-odds assumption is satisfied, the cOR and its 95% CI will be reported; If the proportional odds assumptions are not satisfied, win ratio method will be presented as a supplementary exploratory analysis. Safety analyses will utilize the Safety Analysis Set (participants receiving treatment with ≥1 safety assessment). Safety analyses included descriptive statistics for three safety outcomes and assessment of between-group differences. For the primary safety outcome sICH and the secondary safety outcome any ICH, modified Poisson regression was used to calculate RRs with 95% CIs. A Cox proportional hazards regression model was applied to derive hazard ratios for mortality, and Kaplan-Meier curves were generated to visualize survival distributions between treatment groups. Survival curves were compared using the log-rank test. All tests will be two-sided, with statistical significance defined as P < 0.05. The analyses will be adjusted for the following prespecified covariates: age, baseline NIHSS score, baseline ASPECTS, occlusion site, and time from last known well to randomisation.

Prespecified subgroup analyses will assess treatment effect heterogeneity across key prognostic variables potentially influencing 90-day mRS outcomes, including: age (<median years vs ≥ median years), sex (male vs female), last known well to randomisation time window (<6h vs ≥6h), baseline stroke severity (NIHSS <16 vs NIHSS≥16), pre-stroke mRS score (0 vs 1), occlusion site (intracranial internal carotid artery vs middle artery M1/M2 segments), postprocedural reperfusion status (mTICI 2b vs mTICI 3), stroke etiology (large artery atherosclerosis vs cardioembolism vs unknown or other), prior intravenous thrombolysis administration (yes vs no), and intraprocedural rescue therapy requirement (yes vs no).

## DISCUSSION

EVT achieves high rates of successful recanalisation in LVO stroke, yet up to half of these patients experience unfavourable functional outcomes.^[1]^ This persistent disability underscores the critical need for adjunctive therapies targeting pathophysiological processes beyond the initial recanalization. Post-reperfusion strategies focused on mitigating microvascular obstruction and reocclusion represent a promising avenue. We propose adjunctive tirofiban infusion following successful EVT based on its distinct mechanism as a rapid-onset, reversible glycoprotein IIb/IIIa inhibitor.

The ATTRACTION study is positioned as the first and largest randomized controlled trial specifically investigating adjunct tirofiban administered immediately after successful EVT in large vessel occlusion patients. The recently published PEARL trial and ANGEL-TNK trial provided valuable initial evidence for pharmacotherapy after EVT, demonstrating that intra-arterial alteplase was associated with a probability of achieving a good functional outcome at 90 days.^[7, 18]^ While positive outcomes from recent trials like PEARL and ANGEL-TNK support the efficacy of sequential therapy following endovascular therapy, contrasting results such as those from the POST-TNK trial and POST-UK trial also exist, potentially related to factors including the type and dosing of the administered agent. ^[5,6]^ Nonetheless, current meta-analyses generally support the conclusion that adjunctive pharmacotherapy can improve reperfusion outcomes^.[9]^ Our study builds upon this concept of adjunct administration, aiming to contribute further evidence within this evolving framework. Our trial investigates tirofiban, an agent with a distinct mechanism of action. As a glycoprotein IIb/IIIa inhibitor, tirofiban targets platelet aggregation—the final common pathway for thrombosis—rather than fibrin lysis. This mechanistic difference may translate into a divergent efficacy and safety balance compared to fibrinolytic therapy. The ongoing ADJUVANT-2 trial (NCT06373042) shares a similar design focus on post-EVT tirofiban but has yet to commence patient recruitment. Similarly, the ANGEL-DRUG trial (NCT07026318) investigates tirofiban specifically in patients with significant residual stenosis (≥50%) without planned angioplasty/stenting, explicitly excluding those with any confirmed cardioembolic source (including atrial fibrillation, valvular disease, intracardiac thrombus, recent MI, cardiomyopathy with EF <30%, etc.). Consequently, ATTRACTION provides pivotal, timely evidence for the broader application of tirofiban in post-EVT care, filling a significant gap while complementary trials targeting specific subgroups are initiated.

A deliberate design consideration in ATTRACTION was the inclusion of patients with potential cardioembolic sources. This decision stems from the rationale that endothelial injury caused by the thrombectomy device and the ongoing risk of reocclusion at the site of recanalization are phenomena relevant to all stroke etiologies. While one study suggested heightened hemorrhagic risk with tirofiban in cardioembolic contexts, ^[19]^ another study with preponderance of cardioembolic patients receiving post-EVT tirofiban demonstrated no significant excess in symptomatic intracranial hemorrhage compared to controls.^[20]^ Furthermore, optimal antiplatelet management immediately following successful EVT in cardioembolic stroke remains undefined. The present analysis will focus primarily on efficacy within the stroke etiology subgroup; a comprehensive evaluation of safety will be the objective of the future dedicated analysis.

While the 90-day follow-up is the primary endpoint, this duration may be insufficient to fully evaluate the potential benefits of sequential tirofiban therapy in sustaining improved reperfusion, effects which might become more pronounced over a longer period. Therefore, upon completion of this core study protocol, we plan to conduct an extended follow-up analysis of the ATTRACTION trial cohort. This analysis will assess the long-term impact of the adjunctive reperfusion strategy on clinical outcomes, including but not limited to sustained functional status (e.g., mRS), recurrent stroke, and other vascular events. We believe data on these long-term outcomes will be of substantial value for guiding clinical practice and informing healthcare policy decisions.

We acknowledge some limitations of the current trial. The study population is exclusively derived from China who achieved successful recanalization, the generalizability of the trial results is limited to this population. Moreover, our study focused on patients with large vessel occlusion stroke achieving successful recanalization. Consequently, these findings cannot be generalized to the broader stroke population, including those with posterior circulation strokes or failed recanalization. Currently, a trial investigating posterior circulation stroke (BASILAR-2, ChiCTR2500101044) is in progress, and we anticipate that its results will complement our trial. The principal strength of ATTRACTION lies in its robust design: powered to detect clinically meaningful treatment effects with a large sample size.

The ATTRACTION trial enrolled its first patient on 9 April 2024 and finish enrollment on September 2025 and complete follow-up by December 2025.

## CONCLUSION

The ATTRACTION trial will provide evidence on the efficacy and safety of sequential tirofiban therapy after successful recanalisation in patients with anterior circulation large vessel occlusion stroke.

## Acknowledgements

We are grateful to all the participants in the ATTRACTION programme for their contributions to this study.

## Contributors

Xiang Luo, Wei Wang and Zhouping Tang designed the study; Hao Huang, Shabei Xu, Guo Li, Yi Zhang and Yiming Luo drafted the manuscript; Qianqian Kong, Yi Xie, Chenchen Liu, Gang Deng, Yihui Wang, Donghui Ao, Lingling Lan and Ying Yu provided critical comments/revisions of the manuscript. Xiang Luo is the guarantor.

## Patient consent for publication

Not applicable.

## Ethics approval

This study involves human participants and approved by ethics committee of Tongji Medical College, Huazhong University of Science and Technology (IRB approval number: 2023-S206). All patients or their legal representatives must provide written consent.

## Provenance and peer review

Not commissioned; externally peer reviewed.

## Data availability statement

Data are available upon reasonable request.

