## Supplementary Figure for "Adjunct tirofiban treatment after successful endovascular thrombectomy recanalisation in acute anterior circulation ischemic stroke (ATTRACTION): protocol of a multicenter, prospective, double-blind, randomised trial"

**Supplementary figure** **1 Trial schedule**
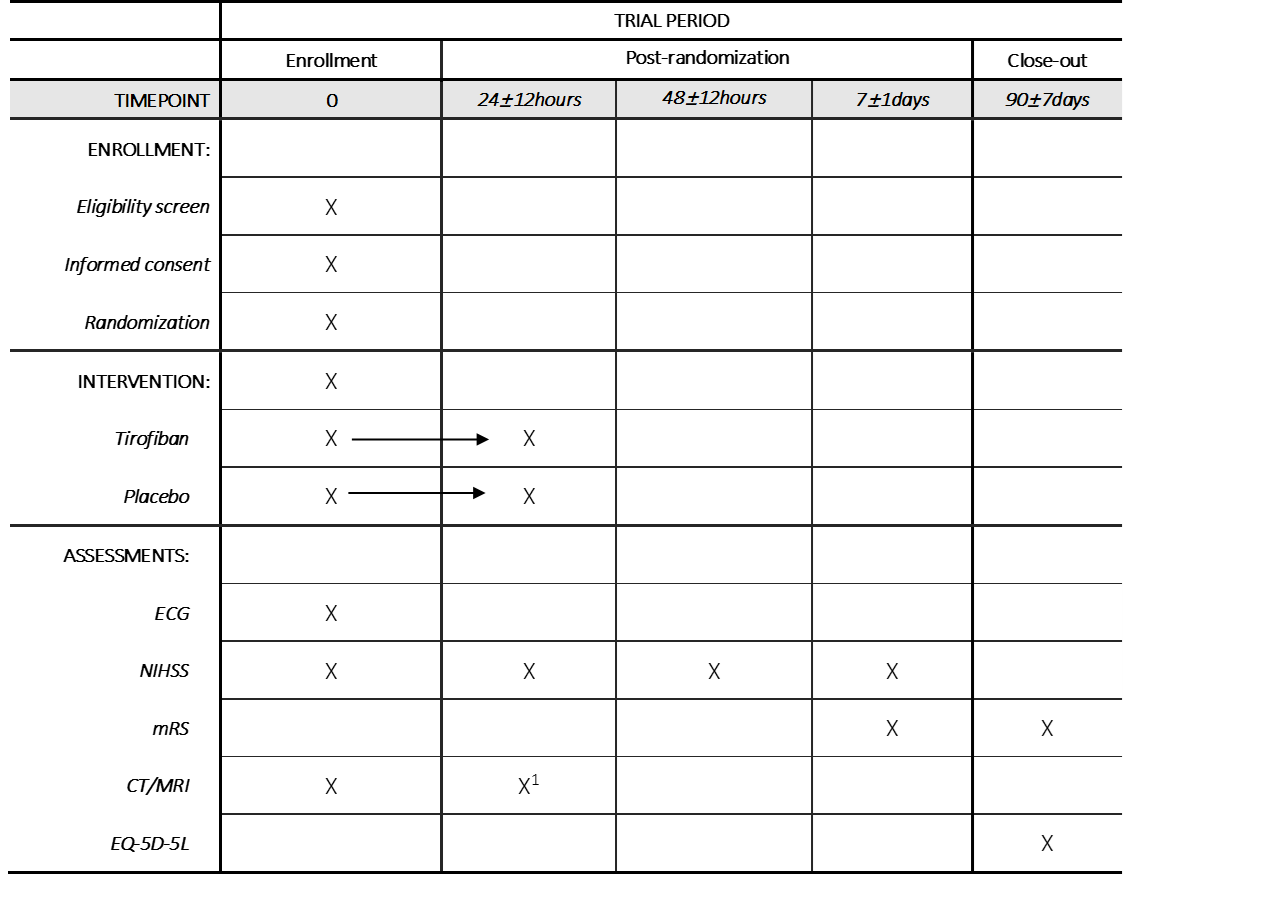

1.CT was repeated at 36 ± 12 hours to exclude cerebral hemorrhage, and MRI was also acceptable if conditions allowed.

ECG, Electrocardiogram; NIHSS, National Institutes of Health Stroke Scale; mRS, modified Rankin Scale; EQ-5D-5L: 5-level 5-dimension health-related quality of life instrument by the EuroQol Group.

**Supplementary figure 2 Distribution map of study sites**
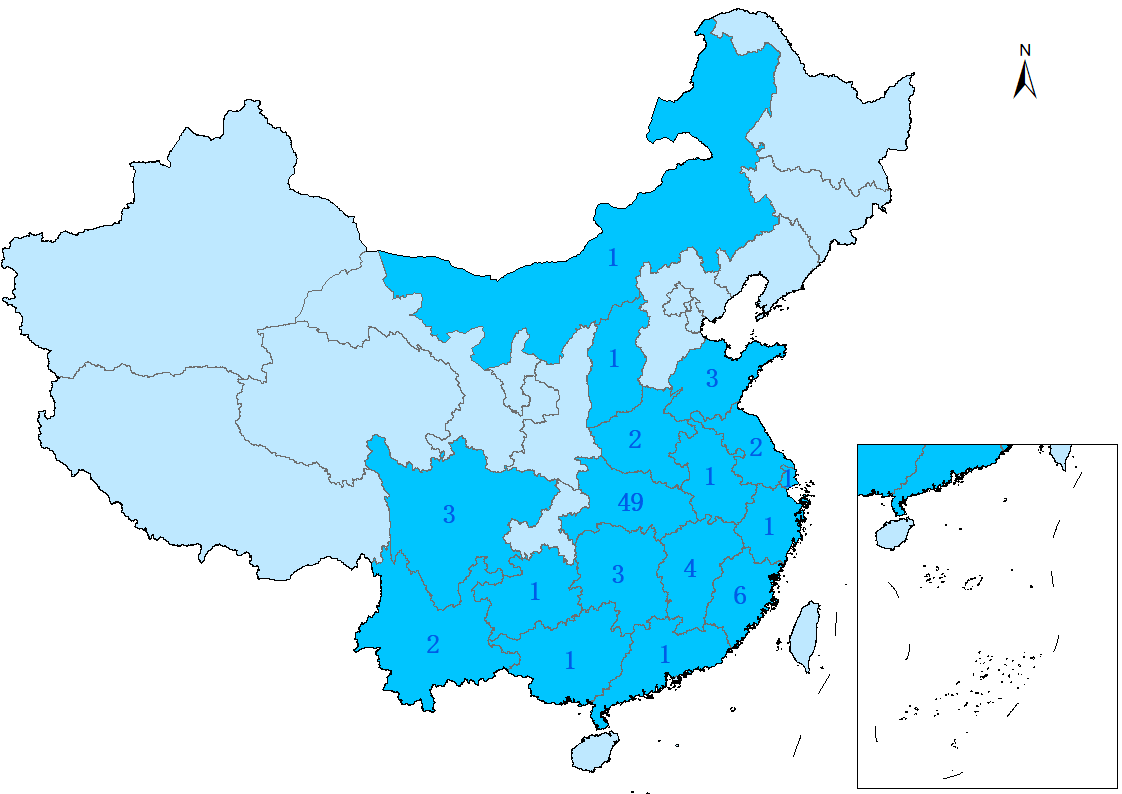

The areas illuminated in blue represent the sites, and the numbers indicate the number of centers.
